# Development and validation of a potential salivary biomarker panel for Oral Squamous Cell Carcinoma

**DOI:** 10.1101/2020.06.08.20125054

**Authors:** Anu Jain, Chinmaya Narayana Kotimoole, Sushmita Ghoshal, Jaimanti Bakshi, Aditi Chatterjee, Thottethodi Subrahmanya Keshav Prasad, Arnab Pal

**Affiliations:** Department of Biochemistry, Post Graduate Institute of Medical Education and Research, Chandigarh, 160012, India; Centre for Systems Biology and Molecular Medicine, Yenepoya Research Centre, Yenepoya (Deemed to be University), Mangalore, 575018, India; Department of Radiotherapy, Post Graduate Institute of Medical Education and Research, Chandigarh, 160012, India; Department of Otolaryngology, Post Graduate Institute of Medical Education and Research, Chandigarh, 160012, India; Institute of Bioinformatics, International Technology Park, Bangalore, 560066, India

**Keywords:** Oral Squamous Cell Carcinoma, Targeted proteomics, Parallel Reaction Monitoring, Biomarker, Early diagnosis, Diagnostic panel

## Abstract

**Purpose:** Oral Squamous Cell Carcinoma (OSCC) is one of the most prevalent cancers in the world with maximum number of cases reported from India. Poor survival rate associated with OSCC can be attributed to non-availability of a biomarker, as one of the major reasons, leading to late presentation. Identification of an early diagnostic biomarker, which can also be used as a screening tool, will be helpful in reducing the disease morbidity and mortality.

**Experimental Design:** In this article we report Parallel Reaction Monitoring (PRM) based validation of 12 candidate proteins, identified initially by TMT tag based relative quantification of salivary proteins on LC-MS, in the saliva of Oral Squamous Cell Carcinoma (OSCC) cases (N=50) and healthy controls (N=49), AZGP1, AHSG, KRT6C, S100A7, S100A9, KLK1, BPIFB2, IGLL5, CORO1A, LACRT, LCN2 and PSAP. Heavy isotope labelled reference peptides were used to produce calibration curve and absolute quantification of proteins and resulting data was analyzed statistically using R.

**Results:** Salivary AHSG (p=*0.0041\*\**) and KRT6C (p=*0.002\*\**) were significantly upregulated in OSCC cases while AZGP1 (p=<*0.0001\*\*\**), KLK1 (p=*0.006\**\*) and BPIFB2 (p=*0.0061\*\**) were significantly downregulated. Multivariate logistic regression modelling resulted in a risk prediction model consisting of AZGP1, AHSG and KRT6C with p value <*0.0001\*\*\**. Using this model ROC with area under the curve of 82.2% was produced and sensitivity and specificity observed for this model was 78% and 73.5%. Positive and negative predictive values for the model were 76% and 75% respectively.

**Conclusion:** We report a potential biomarker panel consisting of proteins AZGP1, AHSG and KRT6C for early diagnosis of OSCC.

## 1. Introduction

Oral cancer, with around 90% cases consisting of squamous cell type, is amongst the top ten prevalent (∼0.6 million) cancers in males around the world [1,2] with approximately 32% cases being reported from India alone. In India, it is the topmost prevalent (∼0.2 million) cancer in males [1,2]. India has an incidence rate of around 0.1 million per year with a mortality rate of 0.07 million. Even with the advancement in the treatment strategies in the last two decades, the survival rate of the disease is still very poor which is often associated with the late presentation of the disease. Non-availability of a suitable tumor marker could be one of the major attributions towards this. Histopathological evaluation of the tumor tissue biopsy along with radiological investigations are the current available diagnostic modality for oral cancer, which is an invasive procedure and advised once the visible symptoms start to appear. The multistep, prolonged and invasive procedure of current confirmatory tool renders it unsuitable as a screening tool. In this context a biomarker will be extremely useful for screening and early detection of the disease [3].

One promising approach to identify the potential biomarkers is to analyse the cancer related proteins in bodily fluids. Saliva, being the potential biofluid for surveillance of general health and diagnosis of disease and in proximity of oral cavity, makes a perfect biological fluid for the identification of biomarker for oral cancer [4,5]. In addition, the non-invasive procedure for collection of saliva makes a salivary biomarker ideal as a screening tool for oral squamous cell carcinoma (OSCC).

In this article we present the results of 12 candidate proteins (identified through Tandem Mass Tag (TMT) based relative quantification of the salivary proteome of OSCC) validated using parallel reaction monitoring (targeted proteomics). Using this approach absolute quantification of the candidate proteins was done in the saliva and data was analysed resulting in a potential biomarker panel as a risk prediction model with high sensitivity and specificity. REporting recommendations for tumour MARKer prognostic studies (REMARK) criteria was followed for reporting the study results [6].

## 2. Materials and methods

### 2.1. Subjects

The study was approval by the Institutional Ethics Committee (INT/IEC/2015/273). A prospective case control study was designed. Patients attending the Department of Radiotherapy and Department of Otolaryngology at Post Graduate Institute of Medical Education and Research, Chandigarh (India), undergoing surgery and/or receiving the standard radio/chemo-therapy with curative intent based on disease stage, decided as per the approved clinical protocol in the institute were enrolled in the study. Fifty, biopsy proven OSCC cases and age and gender matched 49 healthy volunteers were recruited after obtaining informed consent form and following the inclusion and exclusion criteria (supplementary data). Unstimulated saliva samples (at least 5 ml of saliva) were collected, following at least half an hour abstinence from any food and fluid including water, by collecting the saliva directly in a 50mL centrifuge tube. The collected sample was centrifuged at 5000 rpm for 20 minutes at 4°C and supernatant was collected and preserved at -80°C for further analysis. Patients were followed up after treatment completion till the end of the study or till the event (progressive disease) was recorded.

### 2.2. Selection of dysregulated proteins as potential candidate for biomarkers

The candidate proteins were selected from a preliminary shotgun proteomic data obtained by TMT tag based relative quantification of salivary proteins of OSCC cases on LC-MS (data not presented) where 135 dysregulated salivary proteins (supplementary table 1) were identified. These proteins were analysed for their gene ontology, protein-protein interaction network and fold change to select the candidate proteins. With this strategy, 12 highly dysregulated proteins (table 1), also reported to play significant role in cancer biology were selected for further analysis by Parallel reaction monitoring (PRM) based absolute quantification on mass spectrometer.

**Table 1:**
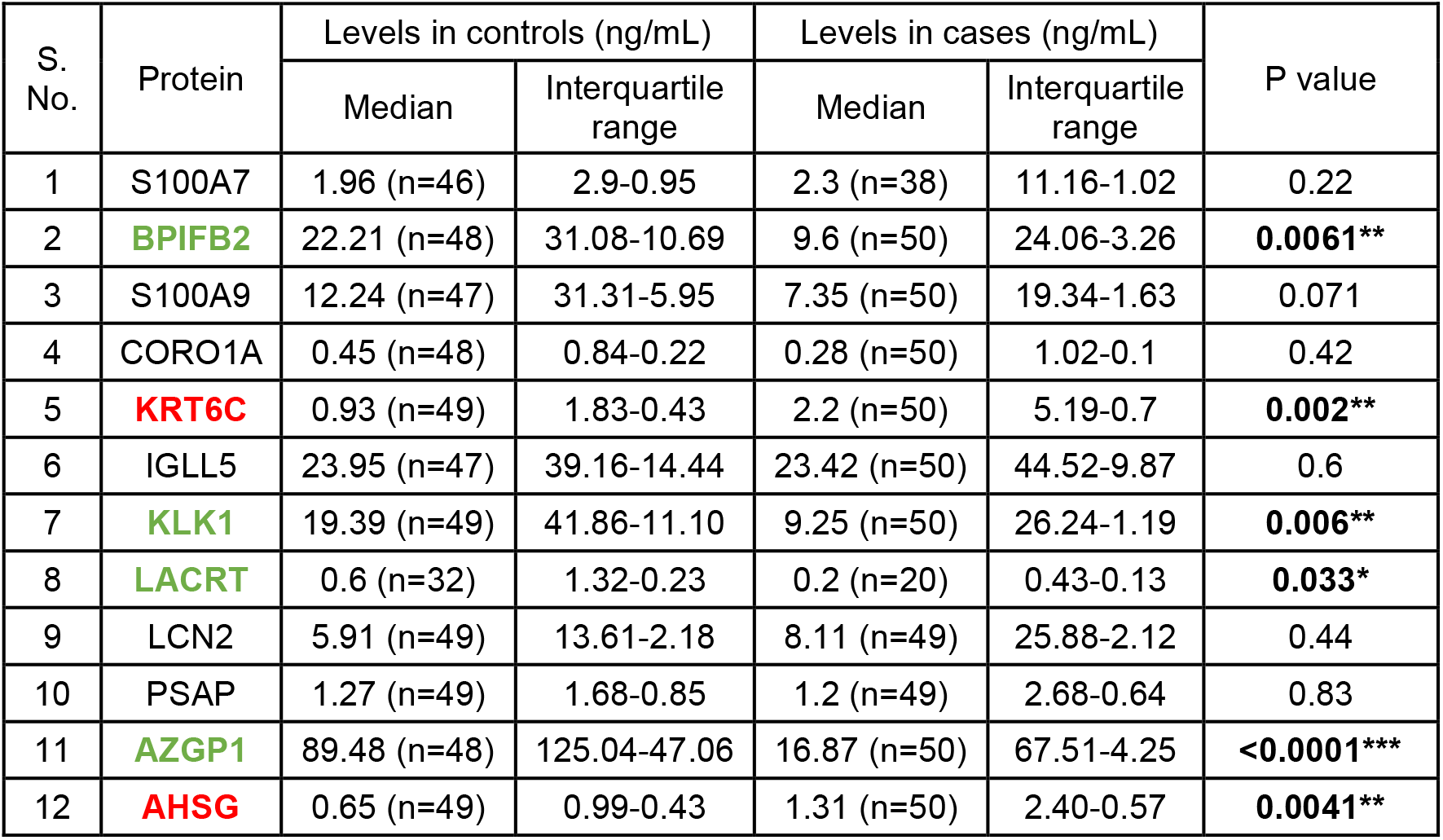
Median levels of protein along with interquartile range as quantified using PRM. p value reported was obtained using Wilcoxon Sum Rank test. Significantly upregulated proteins are highlighted in red and significantly downregulated proteins are highlighted in green.

### 2.3. Standard reference peptides for Parallel Reaction Monitoring (PRM)

Quantotypic unique peptides (supplementary data) were chosen corresponding to the candidate proteins following the selection criteria for peptides for PRM. Tryptic peptides were purchased in the lyophilized form from JPT Peptide Technology (Berlin, Germany) in both light version and labelled version, where C terminal amino acid (lysine or arginine) was heavy labelled (K*= Lys U-^13^C_6_; U-^15^N_2_, R*= Arg U-^13^C_6_; U-^15^N_4_). Peptides were reconstituted as per the manufacturer’s instructions to a final concentration of 100 pmoles/µL and serially diluted ranging from 256 fmol/µL to 0.5 fmol/µL to obtain ten working standard concentrations.

### 2.4. Sample preparation for Parallel Reaction Monitoring

Total protein in the saliva samples was quantified using the Pierce® BCA Protein Assay Kit (#23227, Pierce Biotechnology, Rockford, USA) and following the manufacturer’s protocol. 50 µg of total protein from each sample was prepared for absolute quantification. Total protein was reduced, alkylated and trypsin digested. Digested samples were desalted using Sep-pak C18 cartridge (Waters), dried and reconstituted at the time of analysis with 0.1% formic acid and spiked in with the heavy labelled peptides with a concentration more than the limit of quantification as determined by the standard curves. (This section is mentioned in details in the supplementary data)

### 2.5. PRM method: sample acquisition and data analysis

PRM method was developed using pool of reference peptide to achieve good resolution and ion abundance. The method development and analysis part are mentioned in the supplementary data in details. Briefly, a 40 minutes liquid chromatography method was developed to resolve the peptides and a two-step mass spectrometer method was set to analyse the eluting peptides. First, a full scan MS was done to identify the precursor masses followed by a targeted MS of the selected precursor ions which were analysed and recorded on the orbitrap analyser. The raw files were imported into the skyline to analyse and obtain the product ion transition area of each peptide precursor. Standard curve with the reference peptide pool was generated to calculate limit of detection and quantification which was used as reference to spike the heavy peptide concentration in the sample digest (Supplementary data). The ratio of light to heavy summed transition area was multiplied with the amount of heavy peptide spiked in for the quantification of respective peptides in the samples. The samples were obtained in triplicate and were averaged for final quantification.

### 2.6. Statistical analysis

R was used for the graphical presentation and statistical analysis of the data [7]. Shapiro Wilk normality test was used to check the distribution of the data. Wilcoxon Sum Rank test was used to compare the median protein levels between two groups. Receiver Operating Characteristic (ROC) curve was generated to find out the optimum sensitivity, specificity and cut-off levels of proteins. Multivariate logistic regression was done to analyse the cumulative diagnostic potential of the proteins.

## 3. Results

### 3.1. Patient demography

Among the recruited cases and controls 80% were males and 20% were females. The mean age of cases and controls was 54.6 years and 54 years respectively. 78% (n=39) cases of the of the total, were diagnosed with late stage disease (TNM stage III/IV) and 22% (n=11) were diagnosed with early stage disease (TNM I/II). Post treatment disease status was recorded for the cases. Only 44 cases could be followed-up to record the status and 6 were lost to follow-up. The status was recorded as NED for the patients having no evidence of disease after treatment completion and progressive disease for the cases having progressive disease. Median follow-up time was 6 months. Ten cases were found to have no evidence of disease after treatment and 34 were having progressive disease (figure 1).

**Figure 1:**
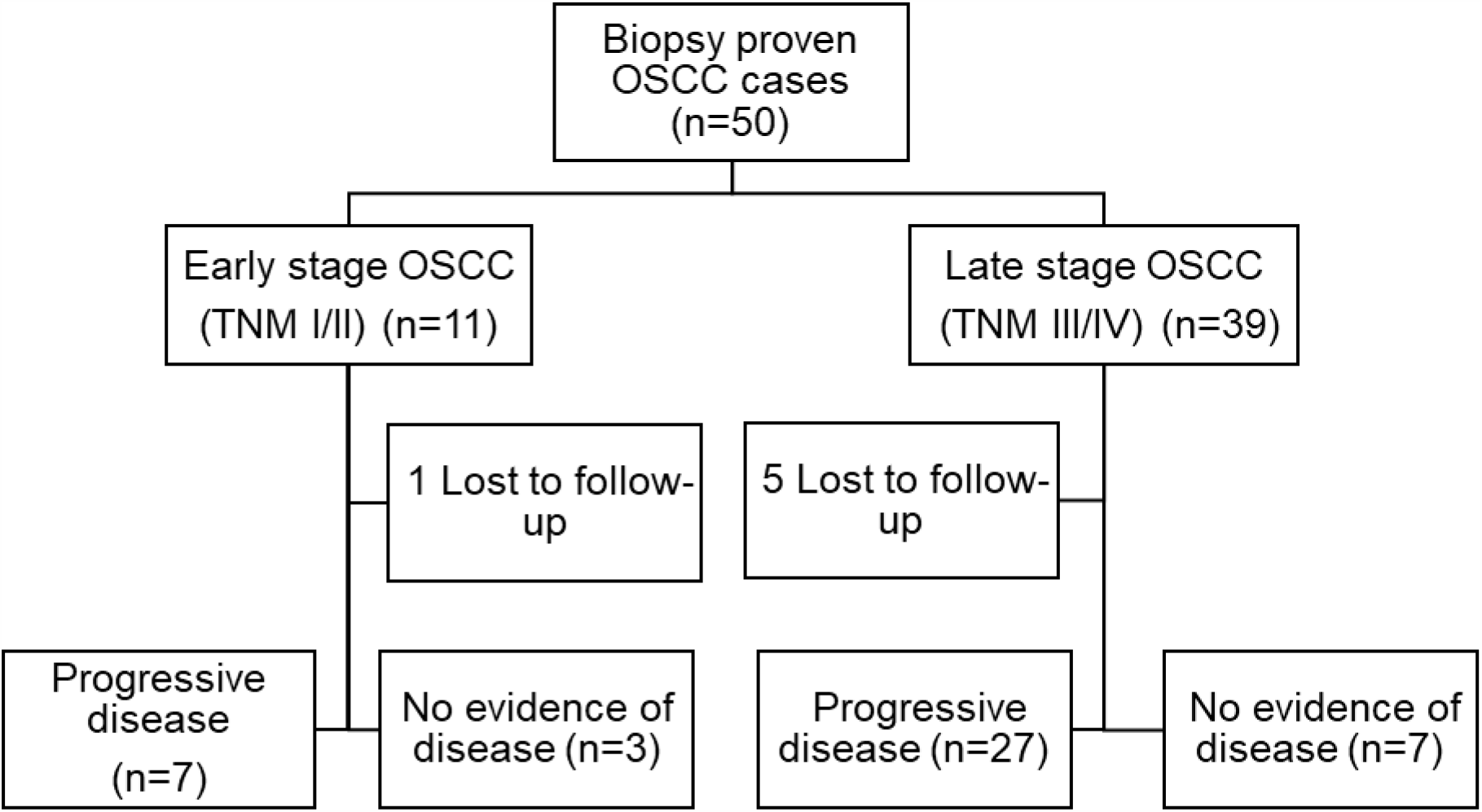
OSCC cases recruited in the study and their outcome.

### 3.2. Five proteins were significantly dysregulated in OSCC cases

The salivary levels of all the 12 candidate proteins is mentioned in table 1. Out of the twelve proteins validated, two proteins AHSG and KRT6C were significantly upregulated and four proteins, AZGP1, KLK1 BPIFB2 and LACRT were found to be significantly downregulated (figure 2) (LACRT was not detected in all the cases and controls so was not included in further analysis).

**Figure 2:**
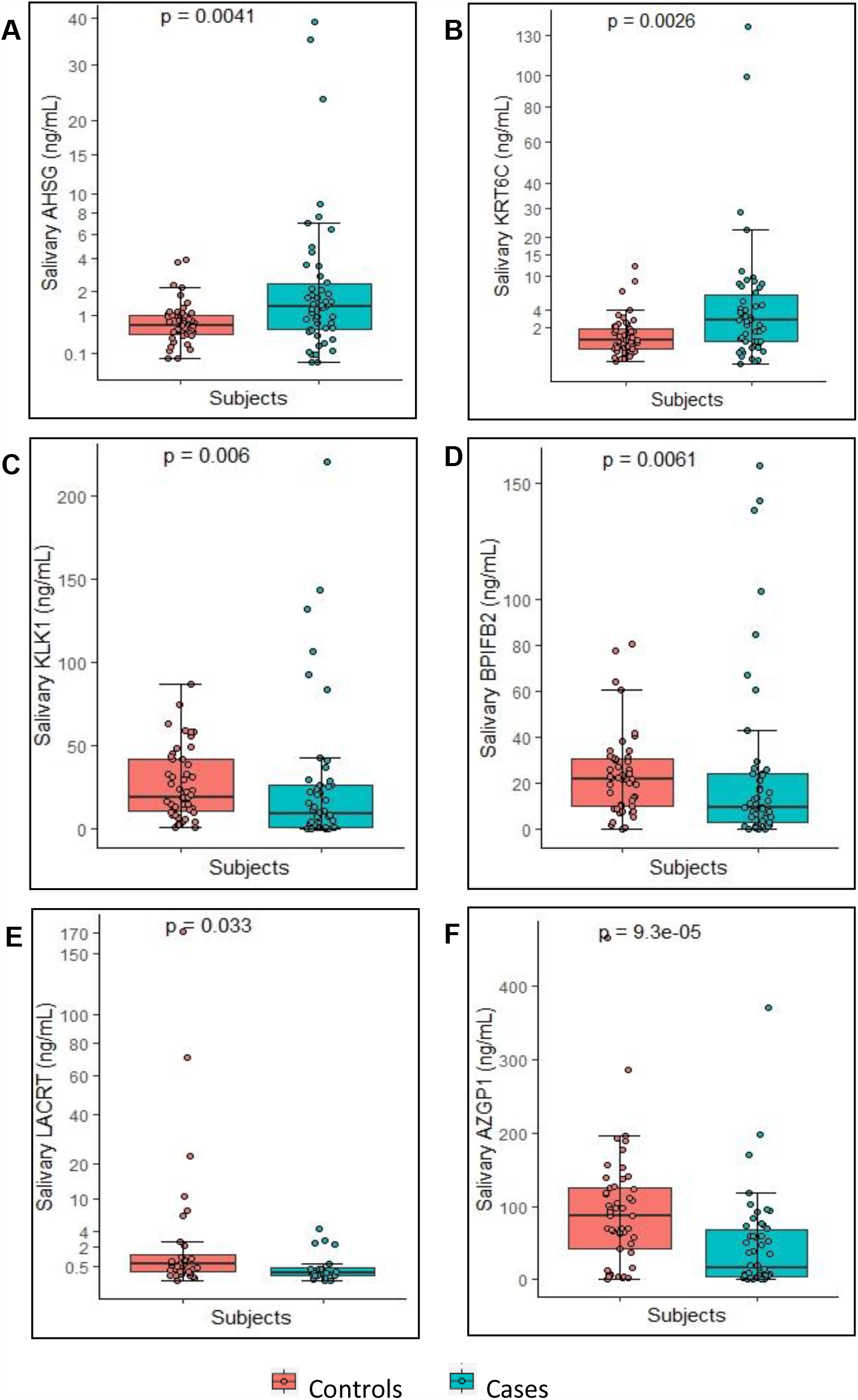
Levels of significantly dysregulated proteins in OSCC. Boxplot representing the levels of dysregulated proteins which were found to be significantly different between OSCC cases and healthy controls (Each dot represents individual value). Proteins **(A)** AHSG and **(B)** KRT6C were significantly upregulated with p value of *<u>0.0041**</u>* and *<u>0.0026**</u>* respectively. Proteins **(C)** KLK1, **(D)** BPIFB2, **(E)** LACRT and **(F)** AZGP1 were downregulated with p value of *<u>0.006**</u>, <u>0.0061**</u>, <u>0.033*</u> and* <u><</u>*<u>0.0001***</u>* respectively. Wilcoxon sum rank test was used to compare the median protein levels of controls and cases (p<0.5 was the cut off for statistical significance). The levels of the proteins are represented in table 1.

The levels of the five significant proteins; AHSG, KRT6C, AZGP1, KLK1 and BPIFB2 were further analysed and compared as per the disease stage (figure 3). It was observed that the levels of AHSG and KRT6C were significantly changed in the late stage disease while for BPIFB2 the difference was significant only in the early stage cases. For AZGP1 and KLK1 the levels were significantly different in both early and late stage disease (Supplementary table 2).

**Table 2:** Characteristics of cumulative ROC curve for proteins AHSG, KRT6C and AZGP1. Area under the curve, p value, sensitivity, specificity positive predictive value (PPV) and negative predictive value (NPV) are mentioned.

| S. No. | Protein | Area under the curve | p value | Sensitivity | Specificity | PPV | NPV | Cut-off |
| --- | --- | --- | --- | --- | --- | --- | --- | --- |
| 1 | Cumulative ROC with proteins AZGP1, AHSG and KRT6C | 82.2% | **<0.0001\*\*\*** | 78% | 73.50% | 76% | 75% | Regression fitted value of 0.502 |
| 2 | Cumulative ROC for early stage | 91% | **<0.0001\*\*\*** | 100% | 72% | 79% | 100% | Regression fitted value of 0.307 |
| 3 | Cumulative ROC for late stage | 84% | **<0.0001\*\*\*** | 82.1% | 76.3% | 78% | 80.6% | Regression fitted value of 0.492 |

**Figure 3:**
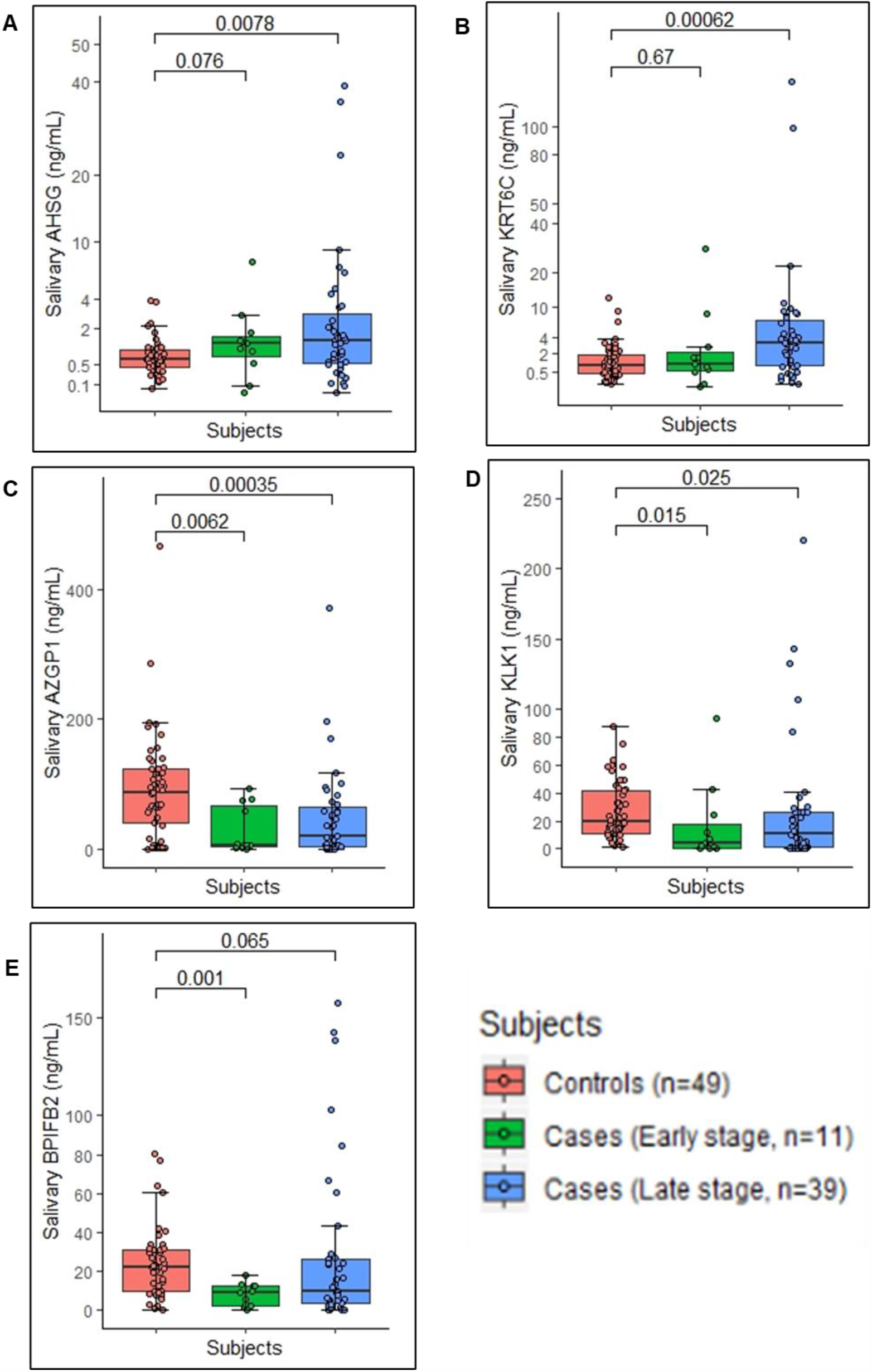
Levels of significantly dysregulated proteins in early and late stage OSCC. Boxplot representing the levels of significantly dysregulated proteins amongst healthy controls, early stage OSCC cases and late stage OSCC cases (Each dot represents individual value). Proteins **(A)** AHSG and **(B)** KRT6C were significantly upregulated in late stage cases. Proteins **(C)** AZGP1 and **(D)** KLK1 were downregulated in both the stages while **(E)** BPIFB2 was significantly downregulated in early stage cases. Kruskal Wallis test was used to check the overall significance.

### 3.3. Sensitivity and specificity of the significant protein

ROC curve was produced using pROC package of R to observe the sensitivity and specificity of the significantly dysregulated proteins. The area under the curve obtained was maximum for AZGP1 (72.8%). The sensitivity and specificity observed were 74% and 74.8% respectively. The area under the curve for AHSG was 66.8% and sensitivity and specificity of 60% and 69.39% respectively. For KRT6C area under the curve was 64% and a sensitivity and specificity of 60% and 63.26% respectively. Area under the curve for both KLK1 and BPIFB2 was 66%. The sensitivity and specificity for KLK1 was 58% and 69.39% respectively and 70% and 64.58% respectively for BPIFB2 (figure 4).

**Figure 4:**
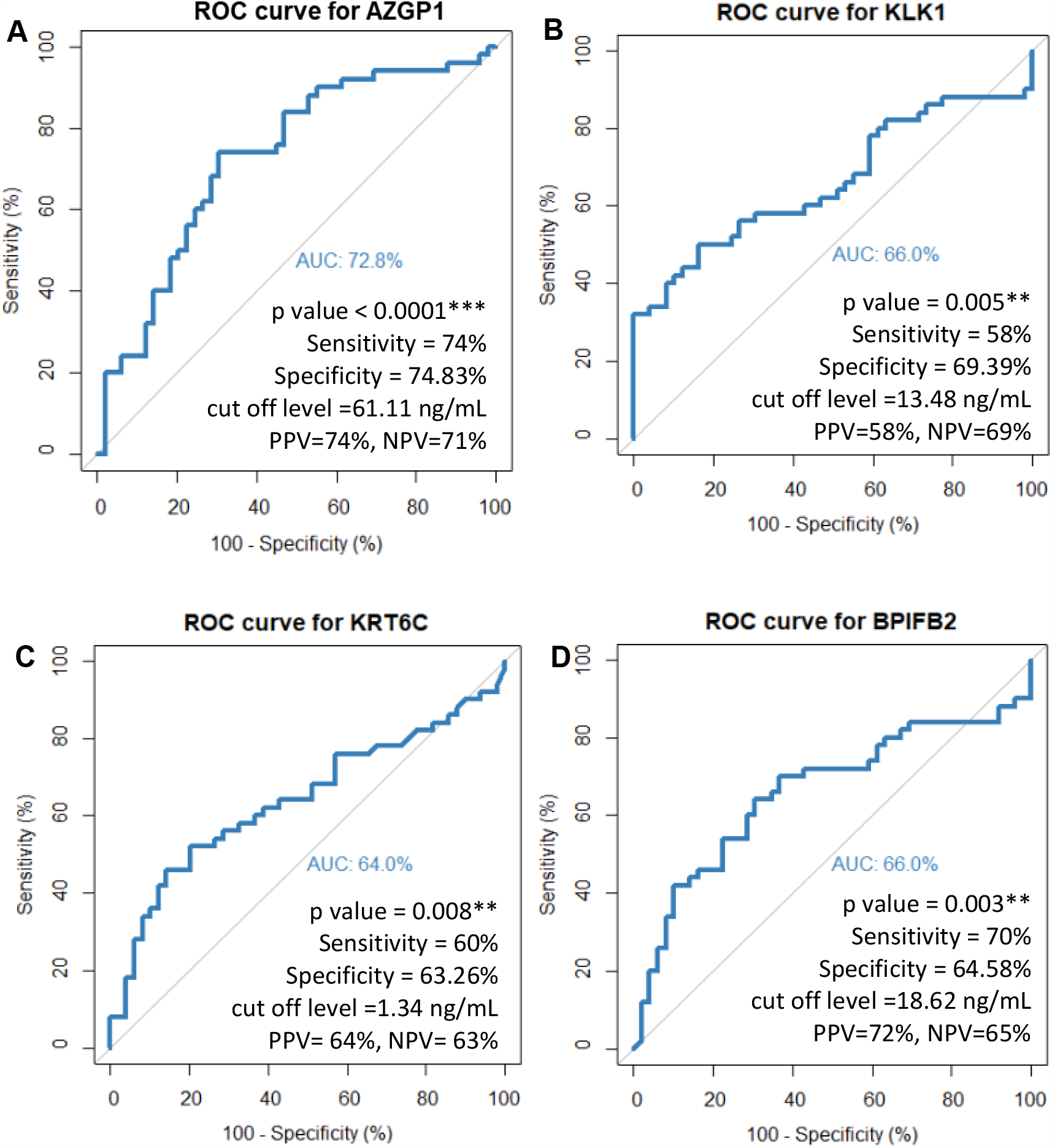
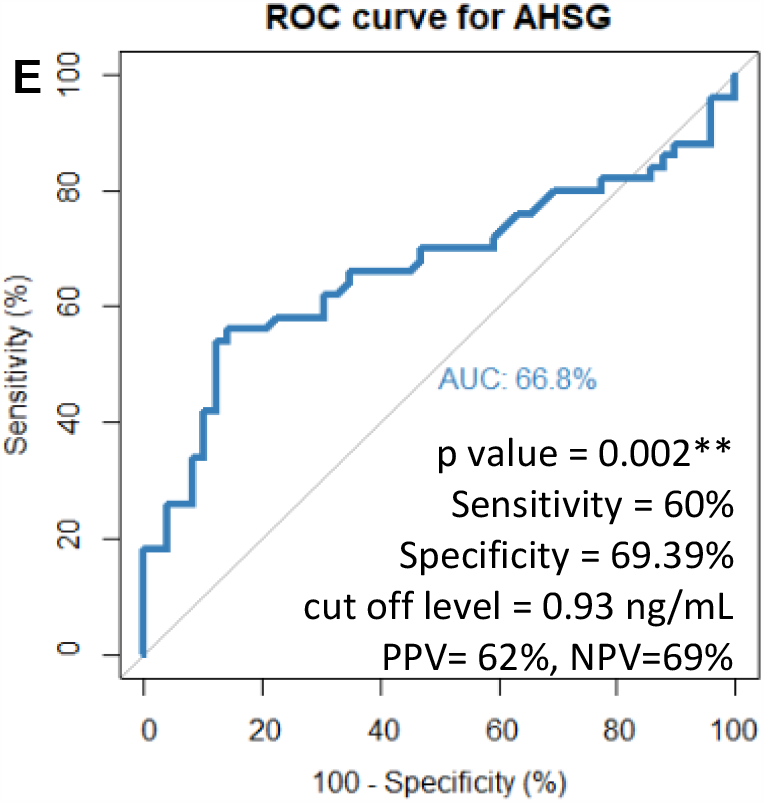
ROC curve for significantly dysregulated proteins. **(A)** AZGP1 **(B)** KLK1 **(C)** KRT6C **(D)** BPIFB2 and **(E)** AHSG. Characteristics of each curve is mentioned in the inset.

### 3.4. AHSG, KRT6C and AZGP1 combinedly make a diagnostic biomarker panel

Univariate logistic regression was applied to check the effect of protein levels on the disease progression. Case and control were selected as the dependent variable and protein levels as the independent variable. AHSG, KRT6C and AZGP1 were found significant upon univariate logistic regression analysis. The beta coefficient for AHSG, KRT6C and AZGP1 was 0.53, 0.184 and -0.01 respectively with a p value *<u>0.012*</u>, <u>0.03*</u>* and *<u>0.003**</u>* respectively. Odds ratio was 1.7 with lower bound of 1.21 and upper bound of 3.21 for AHSG, 1.22 for KRT6C with lower bound of 1.01 and upper bound of 1.52 and 0.98 for AZGP1 with lower bound of 0.97 and upper bound of 0.99. With these significant proteins multivariate logistic regression was applied to obtain a risk prediction model for diagnosis of OSCC. A model with p value <u><</u>*<u>0.0001***</u>* was obtained. Receiver operating curve was obtained for this model. Area under the curve was 82.2% The sensitivity and specificity observed was 78% and 73.5% respectively (figure 5A). Further, the ROC revealed that the model was a little better for early stage OSCC diagnosis (figure 5B) with the area under the curve of 91% for early stage and 84% for late stage.

**Figure 5:**
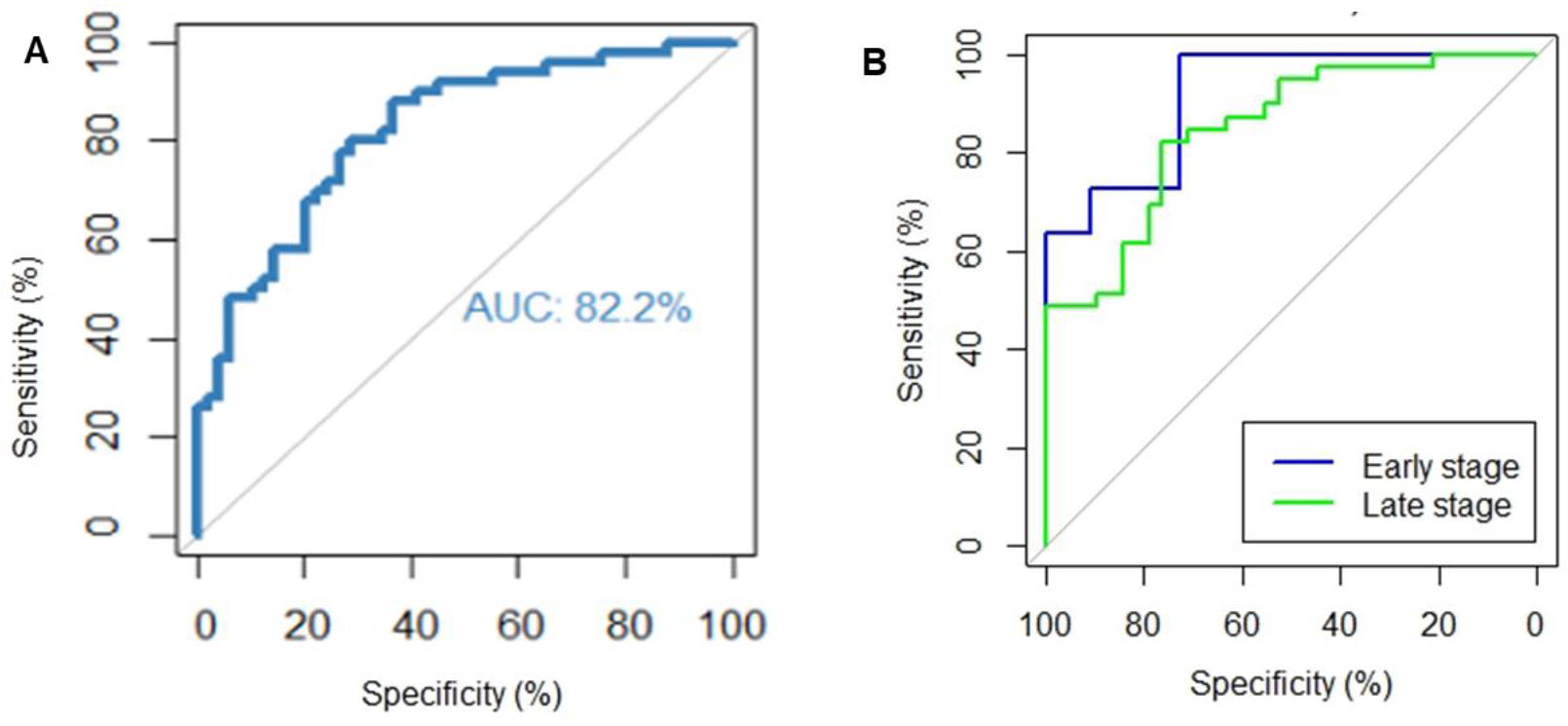
Cumulative ROC curve for AHSG, AZGP1 and KRT6C. **(A)** ROC curve for all the cases and controls **(B)** Cumulative ROC for AHSG, KRT6C and AZGP1 in early and late stage OSCC cases. The characteristics of each ROC curve is mentioned in table 2.

### 3.5. Survival analysis

Kaplan Meier analysis was done to analyse the impact of AHSG, KRT6C and AZGP1 on post treatment disease status of the cases. However, the data obtained was not significant.

## 4. Discussion

Successful identification of a biomarker for early diagnosis of OSCC will improve the overall outcome of the disease and reduce the morbidity associated with the disease, which is currently very dismal. We evaluated salivary proteins as potential biomarker(s) for early diagnosis of the disease which can also be used to screen the high-risk population.

We used parallel reaction monitoring (targeted proteomics) approach which is promising in protein quantification and holds great clinical applications. Parallel reaction monitoring (PRM), where all the transitions are analysed simultaneously in parallel, provides enhanced selectivity with better results, reflected in lower limit of detection and quantification [8]. Using this highly sensitive analytical PRM approach 5 of these 12 proteins were found to be significantly dysregulated in the saliva of our patients.

AZGP1, BPIFB2 and KLK1 were significantly downregulated in our data of which KLK1 and AZGP1 are well reported in cancer but BPIFB2 remains unexplored. AZGP1, an important protein involved in insulin sensitivity and thus play role in metabolism, cell cycle and cancer [9,10]. With metabolic reprogramming as emerging cancer hallmark, AZGP1 expression and role in cancer can be explained. The low mRNA/protein expression of AZGP1 is correlated with disease progression and poor survival in pancreatic cancer [11,12]. Different studies have reported contrasting expression of AZGP1 in different cancers [13–18]. Ibrahim et. al reported low RNA levels of AZGP1 in OSCC tumor tissue of betel quid users [19] which supports our observation at protein level as well. Role of AZGP1 in suppression of cellular invasion and migration [20–22], suggests its association with poor disease response. However, the reduction in levels of AZGP1 in cancer patients mandates a very sensitive detection method for its success as tumor marker in clinical practice. BPIFB2, a member of the lipid transfer/lipopolysaccharide binding protein family. BPIFB2 mRNA expression was reported to be dysregulated in OSCC tumors as compared to the normal counterparts [23] which is in concordance with our observations.

KLK1, a member of serine protease protein family, is involved in a number of physiological functions like remodelling of the extracellular matrix, cellular proliferation and differentiation, angiogenesis, apoptosis etc. The expression of KLK1 was found to be downregulated in a number of cancers including head and neck cancers of which oral cancer is a part [24].

On the other hand, AHSG and KRT6C were significantly upregulated in OSCC. KRT6C, a subtype of type II keratin, has its expression restricted to distinct epithelia type, like filiform papillae of tongue, stratified epithelial lining of oesophagus and oral mucosa and in glandular epithelia [25–27] and associated with abnormal differentiation or enhanced proliferation, like in case of wound healing or cancer with exception of only few body sites [28,29]. The Cancer Genome Atlas (TCGA) also reports high RNA expression of KRT6C in head and neck cancer.

AHSG is a protein of cystatin superfamily with multiorgan expression during embryogenesis [30] which limits to mainly liver and in some cases to osteoblast in human adults [31]. It is a multifunctional protein [32] reported to be associated with various disease conditions [33–36] including cancer [37,38].

However, there are few studies reporting the role of salivary AHSG in disease. To best of our knowledge this is for the first time we are reporting the salivary AHSG levels in oral cancer. In the current study we observed high expression of AHSG in OSCC. The levels were almost twice in cases as compared to the controls as analysed using three different approaches in our laboratory (global proteomics, targeted proteomics and ELISA).

We observed significant difference in AHSG and KRT6C expression between controls and late stage OSCC cases suggesting the role of proteins in disease progression towards aggressive course and this is supported by the reported observations that AHSG is required for the cellular adhesion, proliferation, migration and invasion of the cancer cells. KRT6C expression is also associated with abnormal and enhanced proliferation.

Since cancer is a multifactorial disease, we further used multivariate logistic regression modelling strategy to check the cumulative effect of these proteins as compared to their individual effect. To our expectation, we found that AHSG, KRT6C and AZGP1 together form a highly sensitive prediction model with high sensitivity, specificity and accuracy which is better than their individual diagnostic potential as the average area, sensitivity and specificity of individual proteins increased from 66%, 64% and 67% to 82%, 78% and 73% respectively. This results in a potential biomarker panel for diagnosis of oral cancer with good accuracy. Multivariable biomarker panel approach has been reported to be better in terms of accuracy, sensitivity and specificity, not just for oral cancer but other pathological conditions [39,40]. The panel developed in our study shows better diagnostic accuracy for early stage OSCC but diagnostic potential for late stage OSCC is also considerable. So, as an outcome of this study we report a sensitive biomarker panel which can be developed into a multiparameter rapid testing kit to explore its potential in clinical settings.

For future directions cross validation of this prediction model in terms of accuracy, precision, sensitivity, specificity and positive and negative predictive values using a separate large cohort of cases, disease controls and healthy controls should be done so that the potential value of this prediction model as a biomarker panel in clinical setting can be explored and a rapid detection kit can be developed for the model to facilitate population screening.

## Data Availability

Data will be available upon request from the authors.

## Acknowledgement

This work was supported by Department of Science and Technology-Science and Engineering Research Board (DST-SERB), New Delhi, (EMR/2016/003253) and Post Graduate Institute of Medical Education and Research (PGIMER), Chandigarh, India (71/2-Edu-15/128 & 71/2-Edu-16/4844-45). Indian Council of Medical Research (ICMR), New Delhi, India provided fellowship to Anu Jain [3/1/3/JRF-HRD-022 (10519)]. We acknowledge the logistic help of Prof JS Thakur and Dr. Sudhir Bhandari for collection of control samples and the help of Ms. Kriti, Ms. Rajandeep Kaur, Ms. Deeksha Sachdeva and Ms. Anshika Chauhan for collection and processing of control samples. Contribution of Mrs. Poornima Devadhar and Ms. Anagha Kanichery for sample processing is acknowledged.

## References

[1] Bray F, Ferlay J, Soerjomataram I, Siegel RL, Torre LA, Jemal A. Global cancer statistics 2018: GLOBOCAN estimates of incidence and mortality worldwide for 36 cancers in 185 countries. CA Cancer J Clin 2018;68:394– 424. doi:10.3322/caac.21492.

[2] Ferlay J, Colombet M, Soerjomataram I, Mathers C, Parkin DM, Piñeros M, et al. Estimating the global cancer incidence and mortality in 2018: GLOBOCAN sources and methods. Int J Cancer 2019;144:1941–53. doi:10.1002/ijc.31937.

[3] Nagpal M, Singh S, Singh P, Chauhan P, Zaidi MA. Tumor markers: A diagnostic tool. Natl J Maxillofac Surg 2016;7:17–20. doi:10.4103/0975-5950.196135.

[4] Lee Y-H, Wong DT. Saliva: an emerging biofluid for early detection of diseases. Am J Dent 2009;22:241–8.

[5] Mandel ID. A contemporary view of salivary research. Crit Rev Oral Biol Med 1993;4:599–604. doi:https://doi.org/10.1177/10454411930040034701.

[6] McShane LM, Altman DG, Sauerbrei W, Taube SE, Gion M, Clark GM. REporting recommendations for tumour MARKer prognostic studies (REMARK). Br J Cancer 2005;93:387–91. doi:10.1038/sj.bjc.6602678.

[7] Team RC. The R Project for Statistical Computing. http://www.r-projectorg/ 2013:p1–12.

[8] Gallien S, Bourmaud A, Kim SY, Domon B. Technical considerations for large-scale parallel reaction monitoring analysis. J Proteomics 2014;100:147–59. doi:10.1016/j.jprot.2013.10.029.

[9] Wei X, Liu X, Tan C, Mo L, Wang H, Peng X, et al. Expression and Function of Zinc-α2-Glycoprotein. Neurosci Bull 2019;35:540–50. doi:10.1007/s12264-018-00332-x.

[10] Garrido-Sánchez L, García-Fuentes E, Fernández-García D, Escoté X, Alcaide J, Perez-Martinez P, et al. Zinc-Alpha 2-Glycoprotein Gene Expression in Adipose Tissue Is Related with Insulin Resistance and Lipolytic Genes in Morbidly Obese Patients. PLoS One 2012;7:e33264. doi:10.1371/journal.pone.0033264.

[11] Zhang AY, Grogan JS, Mahon KL, Rasiah K, Sved P, Eisinger DR, et al. A prospective multicentre phase III validation study of AZGP1 as a biomarker in localized prostate cancer. Ann Oncol 2017;28:1903–9. doi:10.1093/annonc/mdx247.

[12] Burdelski C, Kleinhans S, Kluth M, Hube-Magg C, Minner S, Koop C, et al. Reduced AZGP1 expression is an independent predictor of early PSA recurrence and associated with ERG-fusion positive and PTEN deleted prostate cancers. Int J Cancer 2016;138:1199–206. doi:10.1002/ijc.29860.

[13] Parris TZ, Kovács A, Aziz L, Hajizadeh S, Nemes S, Semaan M, et al. Additive effect of the AZGP1, PIP, S100A8 and UBE2C molecular biomarkers improves outcome prediction in breast carcinoma. Int J Cancer 2014;134:1617–29. doi:10.1002/ijc.28497.

[14] Falvella FS, Spinola M, Pignatiello C, Noci S, Conti B, Pastorino U, et al. AZGP1 mRNA levels in normal human lung tissue correlate with lung cancer disease status. Oncogene 2008;27:1650–6. doi:10.1038/sj.onc.1210775.

[15] Ji D, Li M, Zhan T, Yao Y, Shen J, Tian H, et al. Prognostic role of serum AZGP1, PEDF and PRDX2 in colorectal cancer patients. Carcinogenesis 2013;34:1265–72. doi:10.1093/carcin/bgt056.

[16] Huang Y, Li L-Z, Zhang C, Yi C, Liu L-L, Zhou X, et al. Decreased expression of zinc-alpha2-glycoprotein in hepatocellular carcinoma associates with poor prognosis. J Transl Med 2012;10:106. doi:10.1186/1479-5876-10-106.

[17] Huang C, Zhao J, Lv L, Chen Y, Li Y, Jiang S, et al. Decreased Expression of AZGP1 Is Associated with Poor Prognosis in Primary Gastric Cancer. PLoS One 2013;8:e69155. doi:10.1371/journal.pone.0069155.

[18] Tang H, Wu Y, Qin Y, Wang H, Wang L, Guan X, et al. Reduction of AZGP1 predicts poor prognosis in esophageal squamous cell carcinoma patients in Northern China. Onco Targets Ther 2016;2017:85–94. doi:10.2147/OTT.S113932.

[19] Suhr ML, Dysvik B, Bruland O, Warnakulasuriya S, Amaratunga AN, Jonassen I, et al. Gene expression profile of oral squamous cell carcinomas from Sri Lankan betel quid users. Oncol Rep 2007;18:1061–75. doi:10.3892/or.18.5.1061.

[20] Feng M, Feng J, Chen W, Wang W, Wu X, Zhang J, et al. Lipocalin2 suppresses metastasis of colorectal cancer by attenuating NF-κB-dependent activation of snail and epithelial mesenchymal transition. Mol Cancer 2016;15:77. doi:10.1186/s12943-016-0564-9.

[21] Kong B, Michalski CW, Hong X, Valkovskaya N, Rieder S, Abiatari I, et al. AZGP1 is a tumor suppressor in pancreatic cancer inducing mesenchymal-to-epithelial transdifferentiation by inhibiting TGF-β-mediated ERK signaling. Oncogene 2010;29:5146–58. doi:10.1038/onc.2010.258.

[22] Liu J, Han H, Fan Z, El Beaino M, Fang Z, Li S, et al. AZGP1 inhibits soft tissue sarcoma cells invasion and migration. BMC Cancer 2018;18:89. doi:10.1186/s12885-017-3962-5.

[23] Ibrahim S, Jonassen I, Vasstrand E, Amaratunga A, Warnakulasuriya S, Bruland O, et al. Gene expression profile of oral squamous cell carcinomas from Sri Lankan betel quid users. Oncol Rep 2007;18:1061–75. doi:10.3892/or.18.5.1061.

[24] Tailor PD, Kodeboyina SK, Bai S, Patel N, Sharma S, Ratnani A, et al. Diagnostic and prognostic biomarker potential of kallikrein family genes in different cancer types. Oncotarget 2018;9:17876–88. doi:10.18632/oncotarget.24947.

[25] Moll R, Moll I, Wiest W. Changes in the Pattern of Cytokeratin Polypeptides in Epidermis and Hair Follicles During Skin Development in Human Fetuses. Differentiation 1982;23:170–8. doi:10.1111/J.1432-0436.1982.TB01280.X.

[26] Moll R, Krepler R, Franke WW. Complex Cytokeratin Polypeptide Patterns Observed in Certain Human Carcinomas. Differentiation 1982;23:256–69. doi:10.1111/J.1432-0436.1982.TB01291.X.

[27] Oubayoun J-P, Gosselin F, Forest N, Winter S, Franke WW. Cytokeratin patterns of human oral epithelia: Differences in cytokeratin synthesis in gingival epithelium and the adjacent alveolar mucosa. Differentiation 1985;30:123–9. doi:10.1111/J.1432-0436.1985.TB00523.X.

[28] Weiss RA, Eichner R, Sun TT. Monoclonal antibody analysis of keratin expression in epidermal diseases: a 48- and 56-kdalton keratin as molecular markers for hyperproliferative keratinocytes. J Cell Biol 1984;98:1397–406. doi:10.1083/jcb.98.4.1397.

[29] Takahashi K, Paladini RD, Coulombe PA. Cloning and characterization of multiple human genes and cDNAs encoding highly related type II keratin 6 isoforms. J Biol Chem 1995;270:18581–92. doi:10.1074/jbc.270.31.18581.

[30] Dziegielewska KM, Mollgard K, Reynolds ML, Saunders NR. A fetuin-related glycoprotein (?2HS) in human embryonic and fetal development. Cell Tissue Res 1987;248:33–41. doi:10.1007/BF01239959.

[31] Coen G, Ballanti P, Silvestrini G, Mantella D, Manni M, Di Giulio S, et al. Immunohistochemical localization and mRNA expression of matrix Gla protein and fetuin-A in bone biopsies of hemodialysis patients. Virchows Arch 2009;454:263–71. doi:10.1007/s00428-008-0724-4.

[32] Mori K, Emoto M, Inaba M. Fetuin-A: A Multifunctional Protein. Recent Pat Endocr Metab Immune Drug Discov 2011;5:124–46. doi:10.2174/187221411799015372.

[33] Manolakis AC, Christodoulidis G, Kapsoritakis AN, Georgoulias P, Tiaka EK, Oikonomou K, et al. α2-Heremans-schmid glycoprotein (fetuin A) downregulation and its utility in inflammatory bowel disease. World J Gastroenterol 2017;23:437. doi:10.3748/wjg.v23.i3.437.

[34] Majek P, Pecankova K, Cermak J, Gasova Z, Prochazka B, Dyr JE. Alpha-2-HS-glycoprotein plasma level decrease correlates with age in patients with myelodysplastic syndromes. Cancer Biomarkers 2017;20:637–9. doi:10.3233/CBM-170638.

[35] Chen X, Zhang Y, Chen Q, Li Q, Li Y, Ling W. Lower Plasma Fetuin-A Levels Are Associated With a Higher Mortality Risk in Patients With Coronary Artery Disease. Arterioscler Thromb Vasc Biol 2017;37:2213–9. doi:10.1161/ATVBAHA.117.309700.

[36] Nawaz SS, Joy SS, Al Farsi Y, George TP, Siddiqui K. Potential role of serum fetuin-A in relation with pro-inflammatory, chemokine and adhesion molecules in diabetic kidney disease: a case–control study. Mol Biol Rep 2019;46:1239– 46. doi:10.1007/s11033-019-04592-2.

[37] Chen J, Wu W, Chen L, Zhou H, Yang R, Hu L, et al. Profiling the potential tumor markers of pancreatic ductal adenocarcinoma using 2D-DIGE and MALDI-TOF-MS: Up-regulation of Complement C3 and alpha-2-HS-glycoprotein. Pancreatology 2013;13:290–7. doi:10.1016/j.pan.2013.03.010.

[38] Dowling P, Clarke C, Hennessy K, Torralbo-Lopez B, Ballot J, Crown J, et al. Analysis of acute-phase proteins, AHSG, C3, CLI, HP and SAA, reveals distinctive expression patterns associated with breast, colorectal and lung cancer. Int J Cancer 2012;131:911–23. doi:10.1002/ijc.26462.

[39] Lima AR, Pinto J, Azevedo AI, Barros-Silva D, Jerónimo C, Henrique R, et al. Identification of a biomarker panel for improvement of prostate cancer diagnosis by volatile metabolic profiling of urine. Br J Cancer 2019;121:857– 68. doi:10.1038/s41416-019-0585-4.

[40] Dieters-Castator DZ, Rambau PF, Kelemen LE, Siegers GM, Lajoie GA, Postovit LM, et al. Proteomics-derived biomarker panel improves diagnostic precision to classify endometrioid and high-grade serous ovarian carcinoma. Clin Cancer Res 2019;25:4309–19. doi:10.1158/1078-0432.CCR-18-3818.

